# Risk of Heart Failure among Individuals with Metabolic Syndrome Components

**DOI:** 10.1101/2024.06.04.24308459

**Authors:** Rajan Lamichhane, Vijay Bharti, Aarshi R Lamichhane, Kamran Zaheer

## Abstract

Metabolic dysfunction of metabolic syndromes (MetS) has been widely reported to be a significant risk factor for heart failure (HF). While this interaction is well studied, what combinations of MetS factors pose the greatest risk for HF are not well defined. We hypothesize that some components of MetS are of higher risk of HF compared to others. We explored the relationship between the independent components of MetS and their combinatory effect on the risk of HF. All metabolic syndrome components except hypertriglyceridemia were significant individually (P-value<0.001), while hypertension (HTN) and diabetes or insulin resistance (IR) had the higher risk of developing heart failure when taken collectively. The odds of heart failure among the individuals who had HTN and IR was 7.7 times those who didn’t have any MetS components. We observed the additive effect of number of metabolic components on HF, for the individuals who had only one MetS symptom, the odd of HF was 5.4 time compared to those who didn’t have any of MetS symptoms. Similarly, these odds of HF were 6.4 and 7.4 times for those who had at least 2 or at least 3 symptoms respectively. Even though obesity is a significant risk factor for cardiovascular diseases, we found the protective effect of obesity on heart failure, which is an interesting result and needs further investigation.

## Introduction

Cardiovascular disease (CVD) is the leading cause of death in the United States ^1^ and CVD is the most common cause of myocardial infarction (MI). An MI occurs every 40 seconds; every year, about 805,000 people in the United States have an MI ^2^ accounting for 13.4% of all registered deaths in 2018 ^1^.

Metabolic Syndrome (MetS) is associated with numerous adverse health outcomes including CVD ^3-9^. MetS is comprised of five components—insulin resistance (IR), hypertension (HTN), hypertriglyceridemia (HG), low high-density lipoprotein cholesterol (HDL), and obesity ^10^.

Several research studies focused on the relationship between MetS and Heart Failure (HF) ^11-13^, but limited work shows the association between MetS factors and HF. The components of MetS have been linked to an increased risk for HF ^14-16^ ; however, their interactions are not defined. It is not common to find individuals with only a single component of MetS, rather, they present together as a constellation often unique to the individual. It is therefore important to study the simultaneous effect of these components on HF as they overlap among the same group of people. Understanding the risk of HF among each of the MetS components could help direct screening and treatment. In our study, we intend to identify the MetS components and their respective combinations that create the highest risk of developing HF.

Our study is focused on the Appalachian population as this region has a disproportionately higher incidence of HF compared to the rest of the United States ^17^. Also, Appalachia performs worse than the nation in several health indicators. According to the Appalachian Regional Commission report ^18,19^, 7 out of the 10 leading causes of death in the United States, including CVD, are higher in the Appalachian Region compared to the nation.

This study of the rural Appalachian population is important to understand the mechanism of HF among this medically underserved population. The presence of comorbidities, like smoking and obesity, significantly affects the long-term morbidity and mortality of patients with HF ^20^. Population studies have shown that metabolic syndrome increases the risk of developing HF and this effect is mediated by insulin resistance. However, obesity is a key component in metabolic syndrome and a customary partner of insulin resistance because it is protective in patients with established HF, although insulin resistance confers an increased risk of dying from HF. Obesity, a key element of MetS, is often present alongside insulin resistance. In patients with HF, obesity can offer a degree of protection, although insulin resistance is associated with an increased risk of mortality from HF. This phenomenon is called the “obesity paradox” accounting for the complexity of the relationship between HF and MetS ^21^. This paradoxical relationship between MetS and HF should further be investigated to understand the relationship between MetS and HF. Our study aims to investigate this complex relationship between MetS and HF, further breaking down into different individual components of MetS. It is important to understand the role of each of the MetS components to develop customized strategies for HF prevention and treatment. We explore the relationship between MetS and its components with HF to determine which individuals will have a greater need for preventive or therapeutic measures aimed at reducing the risk of HF. Recommendations from this study could help in developing strategies and guidelines based on MetS components to alleviate the burden of HF.

## Study Design and Methods

### Data sources and study design

This study included patients ages 18 and older who received service from the Marshall Health Network between January 2017 and September 2023. The study population was primarily located in Central and North Central Appalachia which includes the western part of West Virginia, the Southern part of Ohio, and the eastern part of Kentucky. Patients were classified as with or without metabolic syndrome if they met at least 3 of the following 5 conditions:

- Insulin Resistance (IR): Average Fasting glucose ≥100 mg/dL, HbA1c >= 5.7 at any point or receiving drug therapy for hyperglycemia, or type 2 diabetes listed as a billing diagnosis or under the patient’s problem list.
- Hypertension (HTN): Blood pressure ≥130/85 mm Hg, receiving drug therapy for hypertension, or hypertension listed as a billing diagnosis or under their problem list.
- Hypertriglyceridemia (HG): Average Triglycerides ≥150 mg/dL, receiving drug therapy for hypertriglyceridemia, or hypertriglyceridemia listed as a billing diagnosis or under their problem list.
- Low High-Density Lipoprotein Cholesterol (HDL): Average HDL-C < 40 mg/dL in men or < 50 mg/dL in women, receiving drug therapy for reduced HDL-C or low HDL-C listed as a billing diagnosis, or under their problem list.
- Obesity: Average BMI > 30, obesity listed as a billing diagnosis, or under their problem list.

This retrospective case-control study has 121 HF cases. The equivalent control subjects were identified by matching age, gender, and heart disease history in the ratio of 1:3 using the nearest neighbor propensity score matching method. The patient data relevant to the study was extracted from Marshall Health Network’s Data Warehouse. The study variables are listed below:

1. Demographic variables – Current Age and Gender
2. Body Mass Index (BMI)
3. Family history of ischemic heart disease (IHD) – patients with a family history of ischemic heart disease and other circulatory diseases.
4. History of MI
5. Social History (Tobacco and Alcohol use) – The use of any kind of tobacco, including smoking, or alcohol during the study period.
6. Inherited syndromes – whether the patient had a history of sickle cell disease, Graves’ disease, or sleep apnea.
7. History of Coronary Artery Disease (CAD)
8. Presence or absence of Insulin Resistance, Hypertension, Low HDL, Hypertriglyceridemia, and Obesity
9. The number of MetS criteria met by the patient
10. Metabolic Syndrome (MetS) – Patients who met 3 or more of the 5 conditions mentioned above in variable 8 were classified as patients with MetS
11. Status of HF

This study was approved by Marshall University IRB (FWA 00002704).

### Statistical Approach

In this retrospective case-control study, we implemented the nearest neighbor propensity score method to match the 121 HF cases with 363 control subjects that did not have HF. The cases were matched by age, gender, and heart disease history.

The descriptive summaries were presented as mean ± SE for the continuous variables and frequency (percent) ± SE for the categorical variables. The odds of HF among individuals who met the criteria for MetS and other components were individually calculated. We investigated how MetS components—IR, low HDL, HTN, and HG—individually and simultaneously impacted the risk of HF by using logistic regression models after adjusting for other risk factors (covariates) of HF. This ensures which components of MetS contributed to the higher risk of HF when taken collectively. Once the important components were identified, we compared the risk of HF among the individuals who had those components and their combinations against the other components. In our analyses, HF status was the outcome variable, while MetS status— comprising HTN, HG, HDL, and combinations of previously identified significant components, each taken separately—served as the primary predictor variables. These results were further validated by running logistic regressions of HF on the number of MetS components to assess for an incremental increase in risk upon adding components. In the analyses, we controlled the other variables: age, gender, history of tobacco and alcohol use, history of CAD, and inherited syndromes.

All statistical analyses were performed using SAS version 9.4 (SAS Institute, Cary, NC). The results were considered statistically significant when the p-values were less than 0.05.

## Results

Table 1 presents the distribution of demographic and other controlled variables together with individual MetS components of 484 individuals. In the study population, the median age was 56 years while the average BMI was 39.9 indicating an obese population, 72.3% of the population was overweight or obese. There were more males (54.7%) than females (45.3%). 70.3% of individuals were younger than 50, 19.4% were between 50 and 65, and 10.3% were older than 65. On average, 2.2 MetS criteria were met, and 44.2% of patients met MetS criteria. Most patients showed obesity, HTN, and IR symptoms, and only 14.1% reported either low HDL or high HG. 49.2% had some history of tobacco or alcohol use, 33.7% had previous MIs, 55.5% had a family history of MI and 31.1% had inherited syndromes associated with HF. As the MetS components occurred collectively, their individual existence was low (table 1). Due to the small sample, we could not estimate the individual effect of these components alone on HF.

**Table 1:**
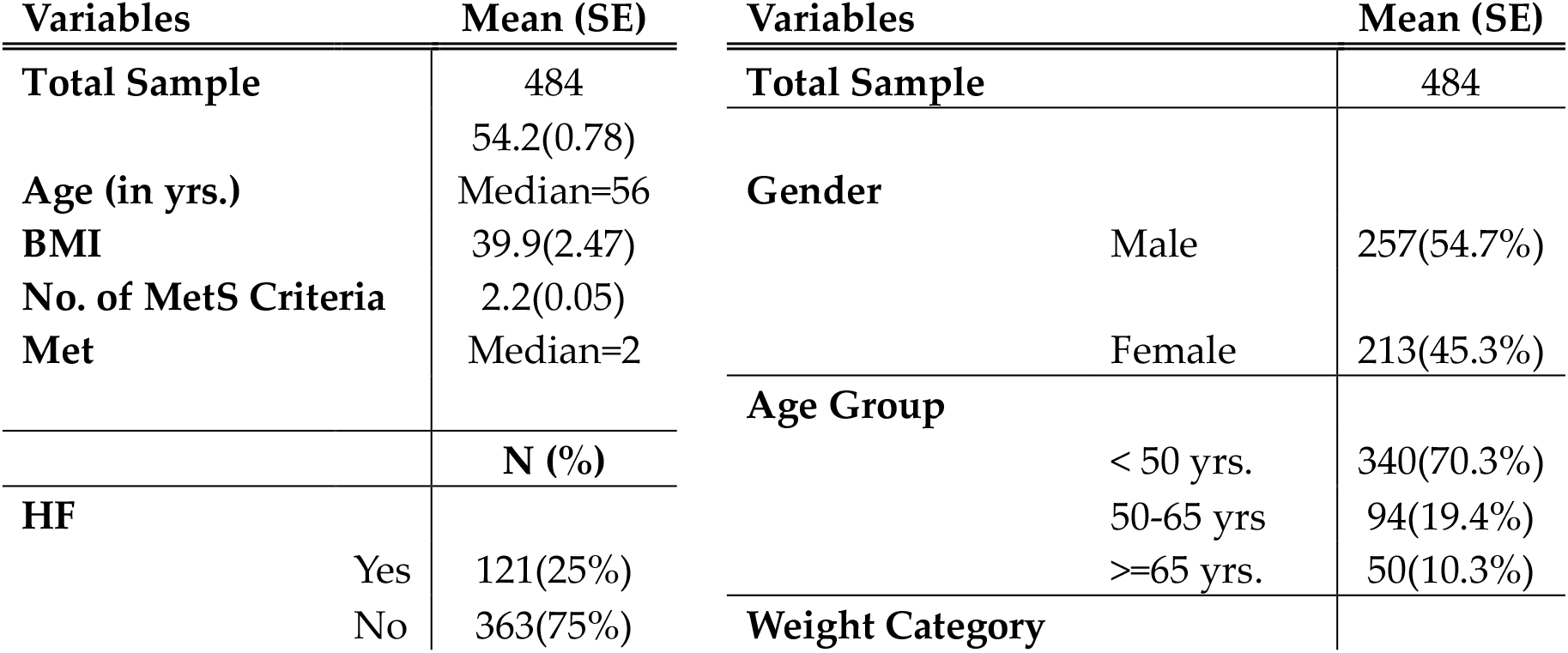
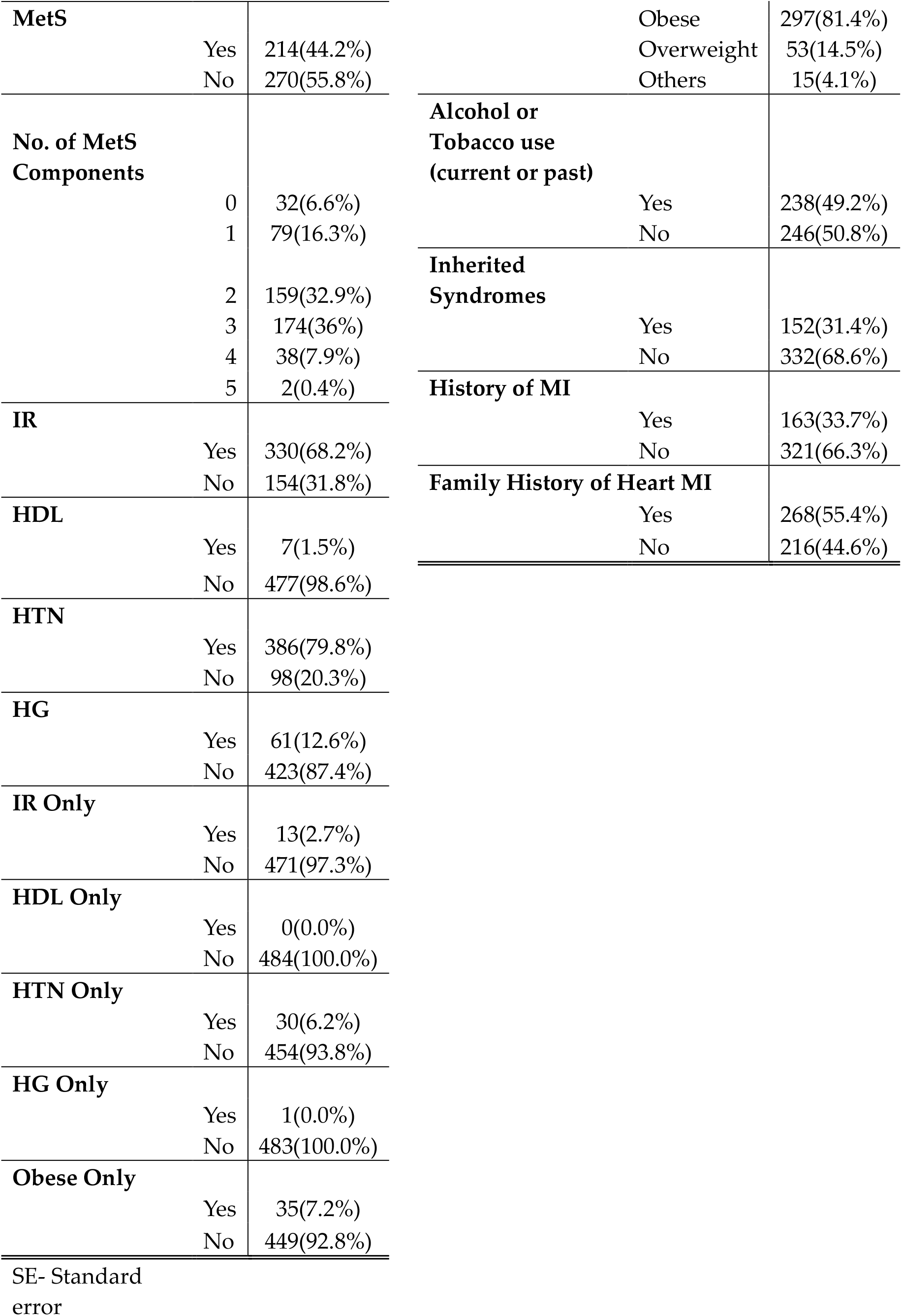
Descriptive Summary.

Table 2a presents the odds of HF among patients with different components of MetS except for low HDL as there was not enough data to evaluate. MetS had a significantly higher risk of HF, and MetS components IR and HTN also had a significantly higher risk of HF. Obesity had some protective effect on HF while the effect of HG was not significant. In all estimations, the covariates mentioned above were controlled to account for their effect on HF status. HF risk is highest among patients with HTN, they had 6.2 times the odds of HF compared to no HTN. Among all patients with HF and HTN, almost 80% were attributable to HTN. Similarly, the odds of HF were 300% higher among patients with IR compared to the patients without, and almost 70% of HF cases in this group were attributable to IR. The odds of HF are twice as high in the MetS group compared to those without MetS. Additionally, the attributable risk of HF due to MetS was 44.3%. Obesity appeared to have a protective effect on HF, the prevention fraction of HF among these individuals was 62.4%.

**Table 2a:** Logistic regression analysis of Heart Failure on different Metabolic Syndrome components.

|  |  | <u>Heart Failure</u> |  | OR (95%<br>CI) | AR% or<br>Prevention<br>Fraction | P-value |
| --- | --- | --- | --- | --- | --- | --- |
|  |  | Yes | No |  |  |  |
| **MetS** |  |  |  |  |  |  |
|  | **Yes** | 71(33.2%) | 143(66.8%) | 2(1.2, 3.1) | 44.3 | 0.005 |
|  | **No** | 50(18.5%) | 220(81.5%) |  |  |  |
| **IR** |  |  |  |  |  |  |
|  | **Yes** | 106(32.1%) | 224(67.9%) | 4(2.2, 7.5) | 69.8 | <0.001 |
|  | **No** | 15(9.7%) | 139(90.3%) |  |  |  |
| **HTN** |  |  |  |  |  |  |
|  | **Yes** | 115(29.8%) | 271(70.2%) | 6.2(2.4, 16.2) | 79.5 | <0.001 |
|  | **No** | 6(6.1%) | 92(93.9%) |  |  |  |
| **HG** |  |  |  |  |  |  |
|  | **Yes** | 15(24.6%) | 46(75.4%) | 0.8(0.4, 1.5) | - | 0.499 |
|  | **No** | 106(25.1%) | 317(74.9%) |  |  |  |
| **HDL\*\*** | **Yes** | 3(42.9%) | 4(57.1%) | NA | 42.4 | NA |
|  | **No** | 118(24.7%) | 359(75.3%) |  |  |  |
| **Overweight/Obese** |  |  |  | 0.2(0.1, 0.6) | 62.4 | 0.004 |
|  | **Yes** | 88(25.1%) | 262(74.9%) |  |  |  |
|  | **No** | 10(66.7%) | 5(33.3%) |  |  |  |

NA- Not enough samples to evaluate

We estimated the effect of more than one component of MetS on HF in Table 2b. We observed that HTN and IR together had a significant risk of HF, the odds of HF among those who had IR and HTN was 7.7 times compared to those who didn’t have any reported MetS components. In all HF cases among the patients who had IR and HTN, almost 82% were attributable to IR and HTN only. The addition of obesity and other factors in the combination of IR and HTN did not increase the odds of HF at a significant rate. Among patients with obesity who also had IR and HTN, the odds of HF were 7.9 times with 81.8% attributable risk compared to those who didn’t have any reported MetS. Due to the observed protective effect of both HG and obesity, the odds of HF among individuals with these components were slightly less than 80%. Also, all other combinations had insufficient data to evaluate the significance.

We justified the above findings by investigating the effect of metabolic syndrome components on HF. Table 3 shows the additive effect of metabolic syndrome symptoms on HF.

**Table 2b:**
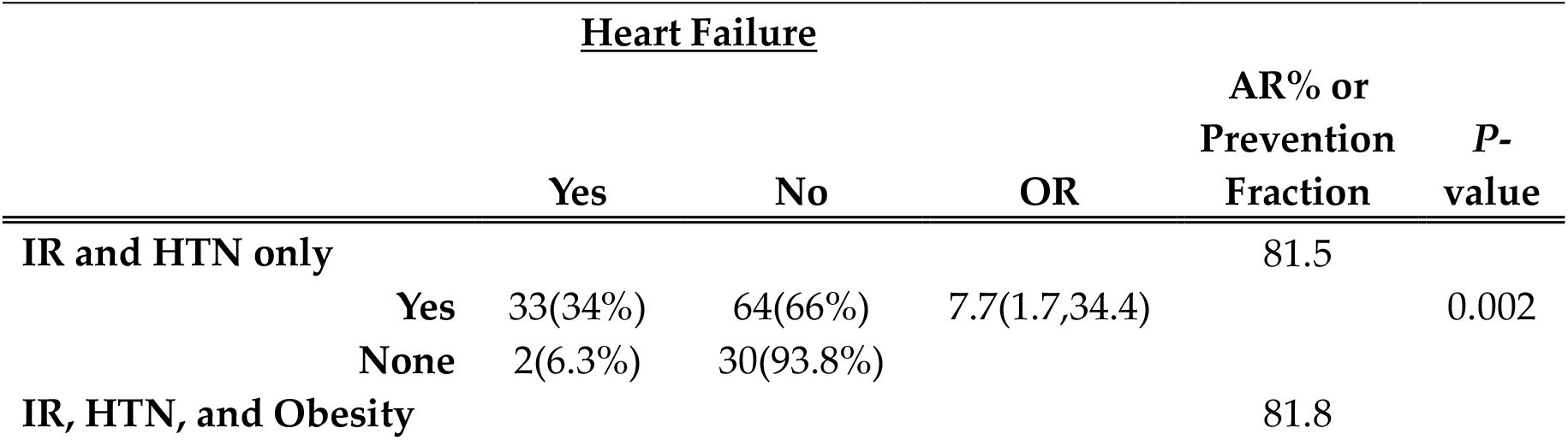
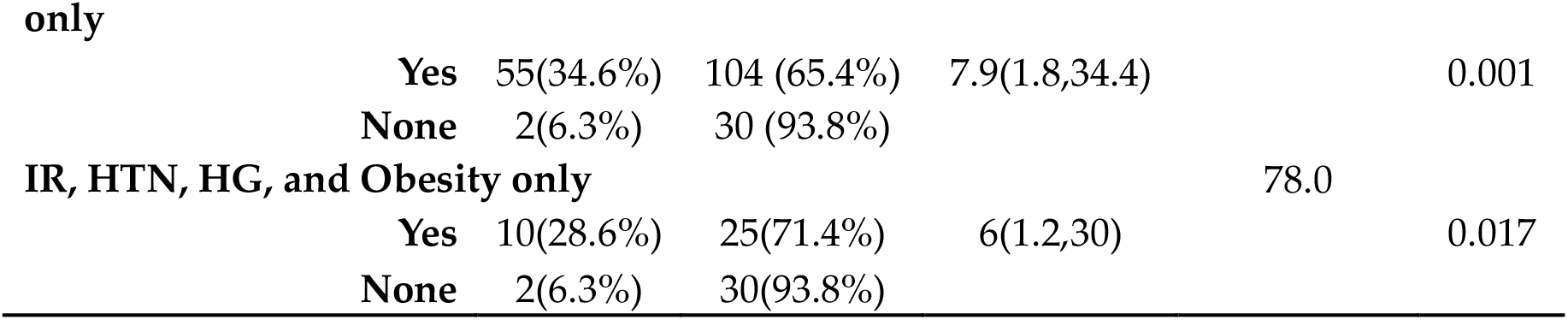
Logistic regression analysis of HF on individual MetS combinations.

**Table 3:** Logistic regression of HF on total number of MetS components.

**Heart Failure**
|  | **Yes** | **No** | **OR (95% CI)** | **AR%** | **P-value** |
| --- | --- | --- | --- | --- | --- |
| **At least one** |  |  |  |  |  |
| **Yes** | 119(26.3%) | 333(73.7%) | 5.4(1.2, 22.8) | 76.0 | 0.023 |
| **None** | 2(6.3%) | 30(93.8%) |  |  |  |
| **At least two** |  |  |  |  |  |
| **Yes** | 111(29.8%) | 262(70.2%) | 6.4(1.2, 27) | 78.9 | 0.012 |
| **None** | 2(6.3%) | 30(93.8%) |  |  |  |
| **At least three** |  |  |  |  |  |
| **Yes** | 71(33.2%) | 143(66.8%) | 7.4(1.2, 32) | 81.0 | 0.007 |
| **None** | 2(6.3%) | 30(93.8%) |  |  |  |
| **At least four\*\*** |  |  |  |  |  |
| **Yes** | 12(30.0%) | 28(70.0%) | 6.4(1.2, 31.3) | 79.0 | 0.021 |
| **None** | 2(6.3%) | 30(93.8%) |  |  |  |

\*\* Not enough data to evaluate

The odds of HF had increased as the number of MetS components increased. The odds of HF with at least one MetS components was 5.4 times (*P* value=0.023) compared to individuals with no components. Similarly, individuals with at least 2 and at least 3 MetS components had odds of HF was 6.4 and 7.4 times, respectively, compared to those with no reported MetS. There were few individuals with at least 4 components; among these individuals, 79% of the risk of HF was solely attributable to at least 4 MetS components.

## Discussion

We explored the relationship between MetS and its components with HF which could provide personalized HF screening, preventive, and treatment options for patients with HF. Overall, insulin resistance and hypertension were the most important components of MetS that could collectively increase the risk of HF. MetS and its components—IR, HTN, and obesity—were significant risk factors for HF. Most patients had multiple components present, so the risk of individual effects could be confounding. Instead, we collectively analyzed the components: the patients with IR and HTN had a higher risk compared to those with at least one other component—HG, obesity, or HTN. There is an increased risk of developing HF as the number of metabolic components increases.

Our findings were in line with past research studies while also generating some novel results. Several studies established the relationship between MetS and HF and indicated the increased HF risk among patients with MetS ^22-23^. Our results also identified MetS as an important risk factor for HF. We identified HTN as the most significant contributing factor to HF risk, which can be correlated to a physiologic basis as follows. Hypertension increases left ventricular afterload and peripheral vascular resistance, leading to pressure and volume-mediated left ventricle remodeling^24^. This remodeling results in diastolic dysfunction, which is often the first sign of increased load. Chronic hypertension, involving both pressure and volume overloads, can lead to either concentric or eccentric left ventricular hypertrophy, associated with heart failure with preserved ejection fraction (HFpEF) and reduced ejection fraction (HFrEF), respectively ^24^.

Diabetes is another important risk factor for HF. IR, the primary endocrine dysfunction of diabetes, disrupts glucose uptake and utilization by cardiomyocytes, leading to energy deficits and impaired contractility^25^. This metabolic inflexibility generates reactive oxygen species and induces mitochondrial dysfunction ^26^. The pathophysiology of diabetes-associated HF involves several key mechanisms. Elevated hemoglobin A1c (HbA1c) levels are associated with an increased risk of HF. Hyperglycemia leads to the formation of advanced glycation end- products and the modification of proteins by β-N-acetylglucosamine, which can negatively impact cardiac function ^27^. Systemic inflammation observed in patients with diabetes likely contributes to cardiac complications. Autonomic dysfunction, disturbances in myocardial metabolism, cardiac remodeling, and poor perfusion are additional mechanisms of diabetes- induced HF^28^.

We were not able to isolate the effects of individual components due to the size limitations as these conditions were rarely observed as a single factor in our study population. However, we found that both HG and low HDL levels were independently associated with an increased risk of HF. Atherogenic dyslipidemia induced by HG and low HDL promotes endothelial dysfunction and atherosclerosis, compromising coronary blood flow and explaining our finding pathologically ^29^.

We observed a protective effect of obesity on HF. This may be due to our choice of measurement unit for obesity-BMI. Individuals with obesity often have preserved skeletal muscle mass and may have a better prognosis secondary to increased stroke volume and thus cardiac output compared with their counterparts who have reduced lean mass and lower stroke volume and cardiac output. Lean mass may play a major role in outcome differences seen in the HF population^30^.

There are several strengths to our study. First, a few studies investigated the relationship between some of the MetS components (IR, HTN) with HF but often did not adjust other risk factors. For most of our analyses, we adjusted for age, sex, BMI, tobacco use, alcohol consumption, family history of HF, and inherited syndromes shown to be related to HF.

Second, it is unlikely to observe just a single component of MetS; it is important to study the simultaneous effect of these components on HF. We investigated the additive effects of metabolic components on HF and compared the risks of HF between patients with MetS and its components. Our study is the first to focus on studying the additive effect of individual metabolic syndrome components.

Third, we performed the risk analysis based on the number of components (table 3) and found the increased odds of HF as the number of components increased, which is a novel finding.

This study also has some limitations. First, the analysis was limited to mainly Central and North Central Appalachia, which may affect the generalizability of the study findings. However, the identified factors reflect the general pattern for the US population. Second, we could not include race, diet, and lifestyle information in the analyses even though they are significant risk factors for HF ^31,32^. There was less racial diversity in the data due to the demographic structure of this region, with more than 90% of the population being White^33^. Future research on HF should consider adding these factors with other factors such as health beliefs, attitude toward HF screening and early detection, lifestyle, and SES as they relate to the disease etiology. Furthermore, we could not perform subgroup analyses by age and gender due to data limitations, so we controlled them in our analyses.

Metabolic syndrome, and its components, insulin resistance, hypertension, and obesity were significant risk factors of HF. Multiple components are often present, and individuals with HTN and IR had a significantly higher risk of HF compared to individuals with no components or any other combinations of components. As the number of MetS components increases, and for individuals with HTN and IR, treatment should be more aggressive for HF prevention compared to those with fewer factors. This will improve the health of our population and combat the marked increase in MetS and HF incidence nationally.

## Data Availability

The deidentified data relevant to this manuscript will be provided upon request.

## Acknowledgment

We acknowledge the Translational Sciences Core of COBRE/ACCORD, Clinical, and Translational Sciences Department, Joan C Edwards School of Medicine, Marshall University for analytics support.

## Non-standard Abbreviations and Acronyms

CVD: CardioVascular Disease
MI: Myocardinal Infaction
MetS: Metabolic Syndrome
IR: Insulin Resistance
HTN: Hypertension
HG: HypertriGlyceridemia
HDL: Low High-Density Lipoprotein
HF: Heart Failure
BMI: Body Mass Index
IHD: Family history of Ischemic Heart Disease
CAD: History of Coronary Artery Disease
OR: Odds Ratio
AR: Attributable Risk
HFpEF: Heart Failure with preserved Ejection Fraction
HFrEF: Heart Failure with reduced Ejection Fraction
HbA1c: Hemoglobin A1c

